# Remote home monitoring (virtual wards) during the COVID-19 pandemic: a systematic review

**DOI:** 10.1101/2020.10.07.20208587

**Authors:** Cecilia Vindrola-Padros, Kelly Elizabeth Singh, Manbinder S Sidhu, Theo Georghiou, Chris Sherlaw-Johnson, Sonila M Tomini, Matthew Inada-Kim, Karen Kirkham, Allison Streetly, Naomi J Fulop

**Author notes:** Corresponding author: Dr Cecilia Vindrola, Senior Research Fellow, Department of Targeted Intervention, University College London (UCL), Charles Bell House, 43-45 Foley Street, London, W1W 7TY, UK.

## Abstract

**Objectives:** The aim of this review was to analyse the implementation and impact of remote home monitoring models (virtual wards) during COVID-19, identifying their main components, processes of implementation, target patient populations, impact on outcomes, costs and lessons learnt.

**Design:** A rapid systematic review to capture an evolving evidence base. We used the Preferred Reporting Items for Systematic Reviews and Meta-Analysis (PRISMA) statement.

**Setting:** The review included models led by primary and secondary care across seven countries.

**Participants:** 27 articles were included in the review.

**Main outcome measures:** Impact of remote home monitoring on virtual length of stay, escalation, emergency department attendance/reattendance, admission/readmission and mortality.

**Results:** The aim of the models was to maintain patients safe in the right setting. Most models were led by secondary care and confirmation of COVID-19 was not required (in most cases). Monitoring was carried via online platforms, paper-based systems with telephone calls or (less frequently) through wearable sensors. Models based on phone calls were considered more inclusive. Patient/carer training was identified as a determining factor of success. We could not reach substantive conclusions regarding patient safety and the identification of early deterioration due to lack of standardised reporting and missing data. Economic analysis was not reported for most of the models and did not go beyond reporting resources used and the amount spent per patient monitored.

**Conclusions:** Future research should focus on staff and patient experiences of care and inequalities in patients’ access to care. Attention needs to be paid to the cost-effectiveness of the models and their sustainability, evaluation of their impact on patient outcomes by using comparators, and the use of risk-stratification tools.

**Protocol registration:** The review protocol was published on PROSPERO (CRD: 42020202888).

**RESEARCH IN CONTEXT:** *Evidence before this study:* Remote home monitoring models for other conditions have been studied, but their adaptation to monitor COVID-19 patients and the analysis of their implementation constitute gaps in research.

*Added value of this study:* The review covers a wide range of remote home monitoring models (pre-hospital as well as step-down wards) implemented in primary and secondary care sectors in eight countries and focuses on their implementation and impact on outcomes (including costs).

*Implications of all the available evidence:* The review provides a rapid overview of an emerging evidence base that can be used to inform changes in policy and practice regarding the home monitoring of patients during COVID-19. Attention needs to be paid to the cost-effectiveness of the models and their sustainability, evaluation of their impact on patient outcomes by using comparators, and the use of risk-stratification tools.

## INTRODUCTION

COVID-19 has rapidly spread across the world, leading to high rates of mortality and unprecedented pressure on healthcare systems. Delays in the presentation of patients with COVID-19 has led to patients arriving as emergencies with very low oxygen saturation, often without accompanying breathlessness (‘silent hypoxia’)^1^. These delayed presentations of severe COVID-19 lead to extended hospital admissions for patients, often requiring invasive treatment and potential admission to intensive care units (ICU) or death^2^. Remote home monitoring models (sometimes referred to as ‘virtual wards’) have been established to: 1) avoid unnecessary hospital admissions (appropriate care at the appropriate place), and 2) escalate cases of deterioration at an earlier stage to avoid invasive ventilation and ICU admission^3^. Some of these models have integrated the use of pulse oximetry to monitor oxygen levels and identify and treat cases of ‘silent hypoxia’^2^.

Remote home monitoring models have been implemented in the US, Australia, Canada, the Netherlands, Ireland, China and UK, with some variation in the frequency of patient monitoring, modality (a combination of telephone or video calls and use of applications or online portals), patient admission criteria, staffing models used for patient monitoring and level of clinical oversight, and use of pulse oximetry^4-8^.

There is a paucity of published literature on the models of care developed to implement remote home monitoring across different healthcare contexts during the COVID-19 pandemic, the experiences of staff implementing these models and patients receiving care, the use of data for monitoring progress, resources required, as well as the impact of these models on clinical, process and economic outcomes. The aim of this review was to address these gaps by identifying the nature and scale of remote home monitoring models implemented during COVID-19, their main components, processes of implementation, target patient populations and lessons learned. We sought to analyse and interpret evaluations of these models and their outcomes.

## METHODS

### Design

We followed the review method proposed by Tricco et al.^9^. The rapid review method follows a systematic review approach but proposes adaptations to some of the steps to reduce the amount of time required to carry out the review. We used a large multidisciplinary team to review abstracts and full texts, and extract data; in lieu of dual screening and selection, a percentage of excluded articles was reviewed by a second reviewer, and software was used for data extraction and synthesis^9^.

We used the Preferred Reporting Items for Systematic Reviews and Meta-Analysis (PRISMA) statement^10^ to guide the reporting of the methods and findings. The review protocol was registered with PROSPERO (CRD: 42020202888, registered 6 August 2020).

### Research questions

The review sought to answer the following questions:

1. What are the aims and designs of remote home monitoring models?
2. What are the main stages involved in delivering remote home monitoring for COVID-19?
3. Which patient populations are considered appropriate for remote monitoring?
4. How is patient deterioration determined and flagged?
5. What are the expected outcomes of implementing remote home monitoring?
6. What is their impact on outcomes and costs?
7. What are the benefits and limitations of implementing these models?

### Search strategy

We used a phased search approach^9^. We carried out a series of search phases where we gradually added search terms based on the keywords used in the literature we identified. Appendix 1 includes the strategies used for each search phase, including the final search strategy. We searched for literature indexed in the following databases: MEDLINE, CINAHL PLUS, EMBASE, TRIP, medRxiv and Web of Science. Initial searches were carried out by CV on 9 July 2020 and updated on 21 August 2020, 21 September 2020 and 5 February 2021. Results were combined into Mendeley and duplicates were removed. The reference lists of included articles were manually screened to identify additional relevant publications.

### Study selection, inclusion and exclusion criteria

One researcher (CVP) screened the articles in the title phase, and additional researchers (KS) cross-checked exclusions in the abstract and full-text phases (KS, MS). Disagreements were discussed until consensus was reached. The inclusion criteria used for study selection was: 1) focus on the monitoring of confirmed or suspected patients with COVID-19), 2) focus on pre-hospital monitoring, monitoring after Emergency Department (ED) presentation and step-down wards for early discharge, 3) focus on monitoring at home (excluding monitoring done while the patient is in healthcare facilities), and 4) published in English. Due to the rapidly expanding evidence-base on COVID-19, we included a wide range of publications (i.e. feature articles, descriptions of services, preprints) and did not limit the selection to evaluations of remote home monitoring.

### Data extraction and management

The included articles were analysed using a data extraction form developed in REDCap (Research Electronic Data Capture) that extracted data on: the design and general characteristics of the model, patient populations, main reported process and clinical outcomes and its potential economic impact. The form was developed after the initial screening of full-text articles. It was then piloted independently by two researchers using a random sample of five articles (CV and KS). Disagreements were discussed until consensus was reached. The data extraction form was finalised based on the findings from the pilot. Data extraction was cross-checked by three researchers (TG, CSJ and ST).

### Data synthesis

Data were exported from REDCap and the main article characteristics were synthesised. The information entered in free text boxes was exported from REDCap and analysed using framework analysis^11^. The initial categories for the framework were informed by our research questions but we were also sensitive to topics emerging from the data.

### Quality assessment

Due to the descriptive nature of the articles and limited data in relation to study design, we did not assess the quality of the studies.

## RESULTS

The initial search yielded 902 articles (Figure 1). These were screened based on the title and abstract and type of article, resulting in 155 articles for full-text review. Full-text review of these articles led to 11 articles that met the inclusion criteria (reasons for exclusion can be found in Figure 1). Three additional articles were identified by reviewing the bibliography, two articles were identified in an updated search carried out on 21 September 2020, and eleven articles were added in an updated search carried out on 5 February 2021, ultimately leading to 27 articles included in the review. We excluded articles that focused on monitoring that took place within hospital settings (i.e. ICU) or for other non-COVID-19 related conditions.

**Figure 1.**
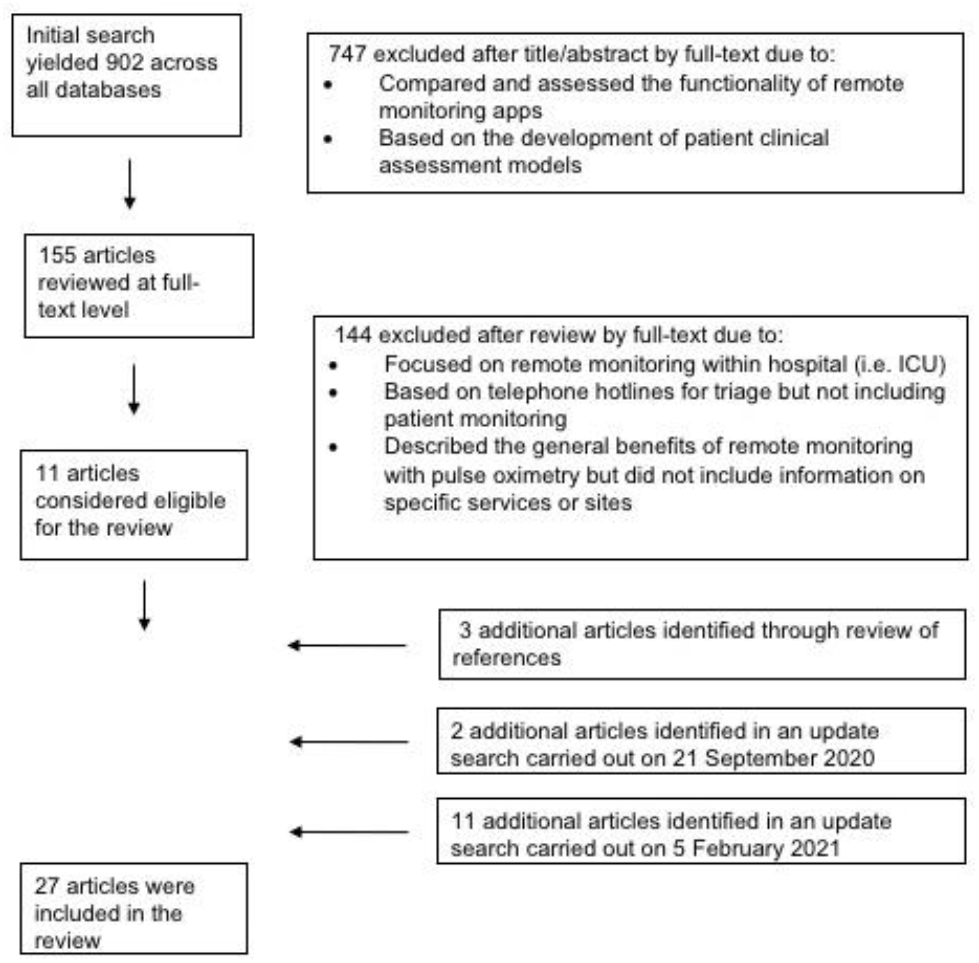
Study selection process

### Characteristics of the included remote home monitoring models

Eleven of the remote home monitoring models were implemented in the US, nine in the UK, two in Canada, two in the Netherlands and one each in China, Ireland, Brazil and Australia. Twelve of the articles described the service, six were identified as evaluations, seven as observational studies, one as a feasibility study and one (containing the example of two models) was a news feature (with a limited description of the services). Eighteen of the examples were published in peer-reviewed journals, nine were published in the form of preprints and one was a published conference abstract. The main characteristics of the included remote home monitoring examples are summarised in Appendix 2.

### Aims and main designs of remote home monitoring models

The primary aim of the remote home monitoring models was to enable the early identification of deterioration for patients self-managing COVID-19 symptoms at home (including those who had not been admitted to hospital as well as those who had been discharged). The programme theory guiding these models was that if patients were able to take the required regular observations whilst remaining at home and communicate these to the healthcare professionals responsible for their care, then cases of deterioration could be identified early and acted upon. These actions could include changing their treatment protocol, referring them to primary care or to the emergency department for assessment and potential admission to hospital. A secondary aim of the models was their use to reduce the rate of hospital infection and demand for beds in the acute care sector, where admission to hospital could be prevented for patients considered suitable to be managed at home and those who had been admitted to hospital could be discharged earlier but continue under the remote care of a medical team (a team that varied in composition depending on the model).

Most of the remote home monitoring models included in the review (23 examples) were led by teams in secondary care. Three examples were primary care led and two were led by both secondary and primary care. Thirteen of the models functioned as pre-admission wards, in the sense that they sought to prevent the admission of patients to hospital or to identify cases of deterioration early (so those who should be referred could be admitted to hospital with lower rates of acuity). Five of the models functioned as “step-down” wards, that is, they were designed for patients who had been admitted to hospital (including ICU) where the medical team had identified that they could be discharged and safely monitored at home until their symptoms improved. Ten models functioned as pre-admission and step-down wards, organised according to two separate pathways.

### Patient populations considered appropriate for remote monitoring

Most of the models established a broad criteria for patient eligibility, defining the patient group as adult (over 18 yrs.) patients with COVID-19 symptoms (suspected and confirmed cases). Six of the models limited referrals to COVID-19 cases confirmed through testing^6 12-15^. The model described by Hutchings et al.^6^ excluded patients over 65 years with significant comorbidities. Shah et al.^16^ excluded pregnant women and only included patients with SpO_2_ above 92% at initial assessment. We did not find any examples targeting socially and economically disadvantaged groups (although some models included support from social workers and mental health professionals)^4 17^. It is important to highlight that the size of the patient cohorts varied considerably (see Table 1 for patient numbers) and ranged from 12 patients to 6853. The models with the highest numbers of patients were implemented in the US. The comorbidities mentioned with greater frequency were hypertension, asthma and obesity.

**Table 1.**
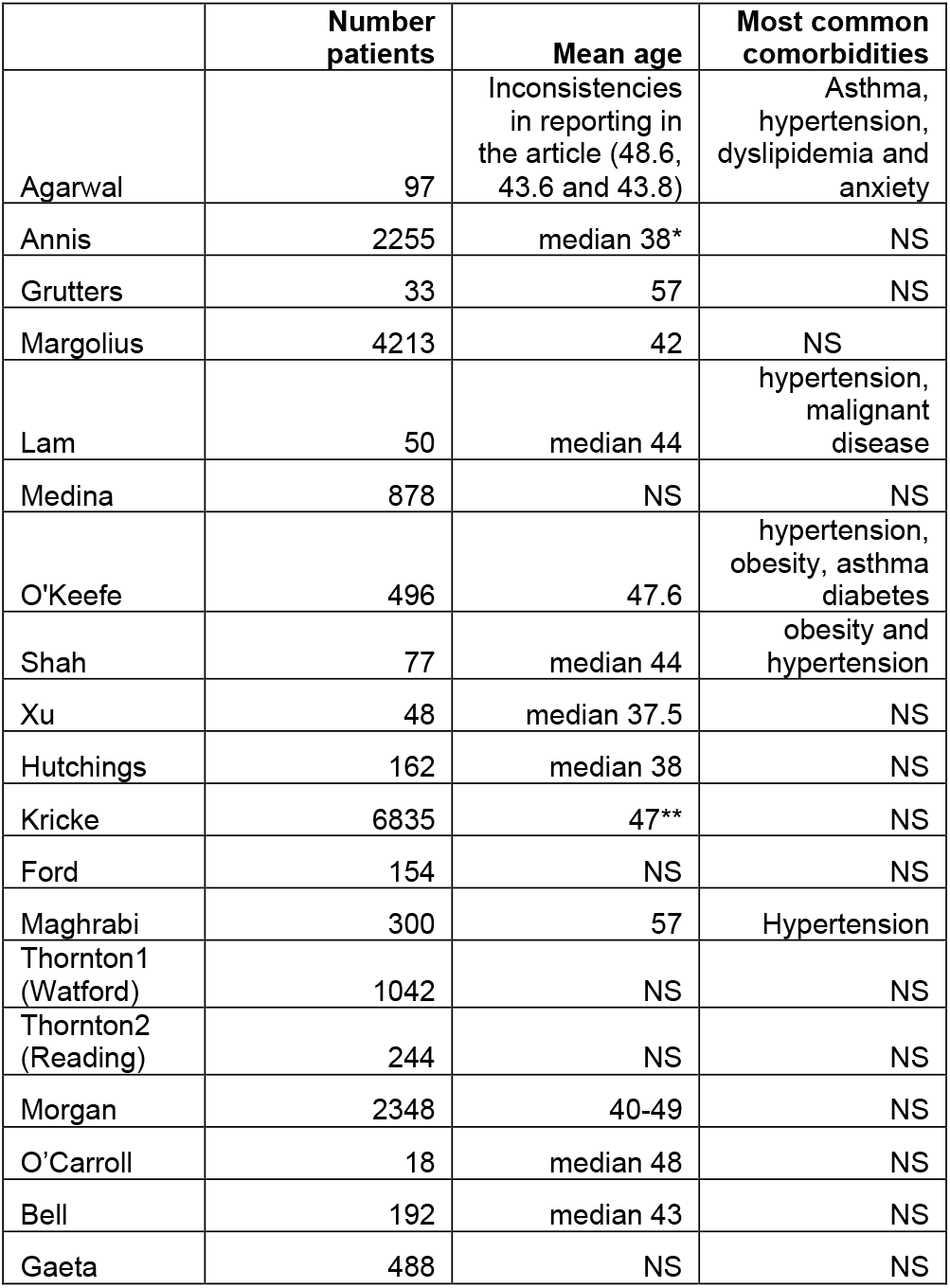
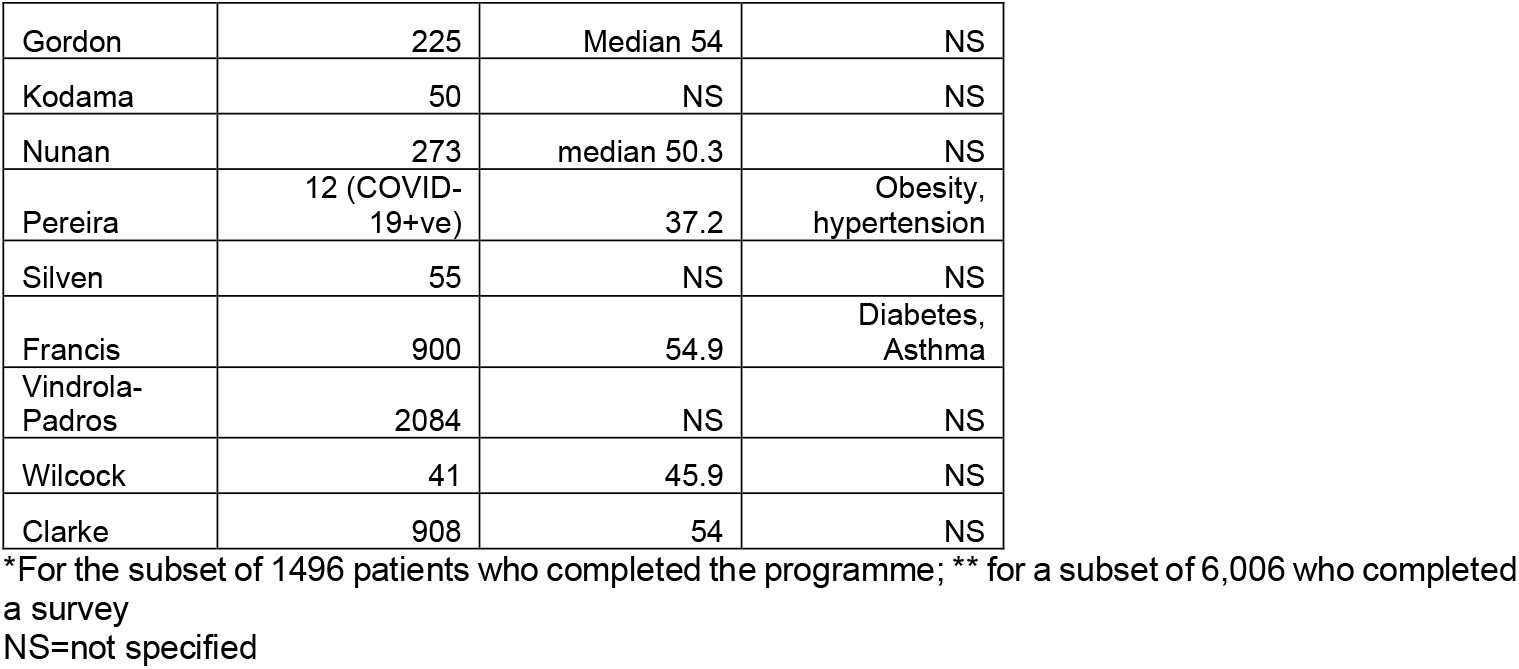
Main characteristics of monitored patients

### Stages of remote home monitoring

The articles described five main stages in remote home monitoring for COVID-19: 1) referral and triage to determine eligibility, 2) onboarding of patient to remote home monitoring service (provision of information to patient and/or carer on monitoring process, mechanisms for escalation and self-care), 3) monitoring (including recording of observations, communication of the information, assessment of the information by the medical team), 4) escalation (if required), and 5) discharge from the pathway.

Patient information recorded at triage included:

- Patient demographics (age, sex, race/ethnicity, insurance type in the models in the US)
- Clinical variables (clinical signs and symptoms, medical history and medications)
- Health data for risk assessment and vital signs data (body temperature, heart rate, respiratory rate and oxygen saturation)

Three studies included some degree of detail in relation to the categorisation of patients in relation to risk^14 17 18 19^. O’Keefe and colleagues^12^ described and evaluated a risk assessment model based on age, medical history and symptom severity. This model was able to identify the need for hospitalisation in initially non-severe COVID-19 patients.

In ten of the examples included in the review, monitoring was based on patient record of observations using a paper-based system and then communicating the information to a member of the medical team by telephone (see Appendix 2). Twelve of the examples relied on the use of an online mechanism, either through an app or online form. Three examples offered patients a telephone or an app option^20^. Another example relied on the use of wearable sensors to continuously monitor temperature readings and transfer these to the medical team^6^. Twenty of the models relied on the use of pulse oximetry from the beginning of implementation, four models did not use pulse oximetry, one model added pulse oximetry three weeks after implementation and two articles indicated that the use of pulse oximetry was being considered in the near future.

Escalation was actioned depending on pre-established thresholds. Not all articles have reported thresholds for escalation and most only refer to the worsening of symptoms. Shah et al.^16^ indicated that patients on their remote home monitoring pathway were flagged as deteriorating if reporting SpO_2_ below 92% after a double reading. Xu et al.^21^ used a SpO_2_ reading of below 93% or BP less than 90/60 mmHg. Some of the examples included in the review established safety-netting options in cases when patients could not be reached via phone such as calling the police so they could visit the patient at home^6^.

Most patients were followed-up until their symptoms improved or the patient opted out of the pathway. Medina et al.^13^ reported following up patients on the step-down pathway for 7 days post-discharge from hospital and those on the pre-admission pathway for 14 days. Shah et al.^16^ followed-up patients on their pre-admission pathway for 7 days. Hutchings et al.^6^ referred patients to their GP for follow-up after discharging them from the remote home monitoring pathway.

### Expected outcomes of implementing remote home monitoring

The outcomes recorded in each remote home monitoring model are listed in Appendix 2. They can be grouped in three main categories: 1) process outcomes related to the remote home monitoring pathway, 2) process outcomes related to secondary care and 3) patient outcomes (including clinical and experience). Process outcomes related to the remote home monitoring pathway included: time from swab to assessment, time to escalation and ambulance attendance/emergency activation (i.e. calling 999 or 911). Process outcomes related to secondary care included length of stay. Outcomes considered at the patient level included: emergency department attendance/reattendance, hospital admission, ICU admission, readmission, mortality, ventilation or non-invasive ventilation needs, and patient satisfaction.

### Impact on outcomes

It was difficult to carry out an analysis of the impact of remote home monitoring across all examples because not all articles reported data on the same outcomes (Table 2). Mortality rates were low, admission or readmission rates ranged from 0 to 29%, and ED attendance or reattendance ranged from 4 to 36%. Six of the models reported data on patient feedback, with high satisfaction rates^5 8 18 22 23 24 25 26^.

**Table 2.** Impact of remote home monitoring on selected outcomes

|  | virtual LoS | Escalation | ED attendance/<br>reattendance | Admission/<br>readmission | Mortality |
| --- | --- | --- | --- | --- | --- |
| Agarwal | 8 days (median) | 5.10% | 4.2% | 0 | NS |
| Annis | NS | NS | 4.0% | 0.6% | NS |
| Grutters* | 13 days (mean) | 18 patients<br>reassessed<br>in hospital | NS | 9%** | 0 |
| Margolius* | NS | NS | 7% | 1% | NS |
| Lam | 12.5 days (only<br>for 52% of<br>patients) | 12% | NS | 8% | 0 |
| Medina | NS | 10% | NS | 2%, 3%** | 9 patients but<br>denominator is<br>unclear |
| O'Keefe | 13.1 days | NS | NS | 7.1% | NS |
| Shah | NS | 25% | 36% | 29% | 2.6% |
| Xu | NS | NS | NS | NS | 0 |
| Hutchings | 8 days (only for<br>62 of the patients in the sample) | 5 patients | 2.5% | 1.9% | 0 |
| Kricke | NS | NS | 7.7% | NS | NS |
| Ford | NS | 14.3% referred to physician review; 3.9% physician to patient call; 2.6% to ED and admitted | 2.6% | 2.6% | NS |
| Maghrabi | 3.5 days (median) | NS | 13% | 9%** | 0.66% |
| Thornton1 (Watford) | NS | NS | NS | NS | NS |
| Thornton2 (Reading) | NS | NS | 11.9% | 7.4% | 0 |
| Morgan | 12.7 days (mean) | 16.9% escalated to nurse review | 7.9% | 3.4% | NS |
| O'Carroll | 12 days (median) | NS | NS | 4 patients | NS |
| Bell | NS | NS | 16.7% | 3.6% | 0 |
| Gaeta | NS | NS | 18.4% | 8.8% | 1.2% |
| Gordon | unclear | NS | 4.9% | 1.3% | NS |
| Kodama | NS | 26% | 6%**** | 2%**** | NS |
| Nunan | NS | NS | 11.4% | 7.0% | 0.4% |
| Pereira | NS | NS | NS | NS | NS |
| Silven | NS | NS | NS | 9% | 0 |
| Francis | NS | NS | NS | 8.1% | 2.0% |
| Vindrola-Padros | NS | 10.4% | 8.3% | 6.4% | 1.1% |
| Wilcock | 10.3 days (mean) | NS | NS | 7.3% | 1.9% *** |
| Clarke | NS | NS | 5.7% | 4.4% | 3.1% |

\*included data for patients on remote home monitoring pathway, \*\*refers to readmission in cases of step-down, wards, \*\*\*of 52 initially recruited, \*\*\*\* these refer to very low patient numbers. LoS=length of stay, ED= emergency department, NS= not specified or not able to calculate based on data reported in the manuscript.

Remote home monitoring process outcomes were only included in six of the articles, with time from swab to assessment ranging from 2 to 3.7 days^12 17 20^ and virtual length of stay from 3.5 days to 13 days (see Table 2). Only one article presented findings on reduction in length of stay, calculated at 5 days fewer per patient^26^.

### The economic impact

Very few of the selected studies for this rapid review provided a descriptive form of economic analysis, though some of them mentioned the potential for cost savings based on the utilisation of virtual monitoring programs for other treatments in similar settings^14 22 26^. The study by Nunan and colleagues^22^ found that setting up a remote oximetry monitoring at the Royal Berkshire Hospital resulted in cost avoidance (in terms of bed days, saturation probes and staffing wages) that amounted to £107,600 per month. The amount spent per patient on remote monitoring varied by country and type of costs included in the analysis. The study from Gaeta and colleagues^27^ reported a total cost of $621.8K (equivalent of £485.0K using purchasing power parity) for 621 COVID-19 patients that were monitored using outpatient telehealth follow-up in the Brooklyn Methodist Hospital. These costs included also costs of inpatient follow-up and averaged at £781.0 per monitored patient, whereas the mean cost per monitored patient reported in England varied from £400 to £553 for step-down and pre-hospital models respectively^24^. Some of the selected studies highlighted the fact that, during the pandemic, the intervention used existing resources and staff that were made available due to the emergency situation^7 12 14 28^. However, they also highlighted that, with the return to normal workloads in the health care system, a question of allocation of resources and sufficient staffing still remains.

## DISCUSSION

In this article we have sought to make a contribution to the rapidly growing evidence-base on the use of remote home monitoring models for patients with confirmed or suspected COVID-19 (see box 1 for key lessons). The review has pointed to factors that need to be taken into consideration in relation to the design of these models. Most of the models included in the study were led by secondary care but some authors argued that coordination between primary and secondary care could facilitate the implementation of remote home monitoring pathways^5 7 13^. Primary care led models might be more adaptable to evolving patient and system needs and easier to replicate in contexts with limited secondary care access and capacity^17^. Three models integrated mental health and social care support during and after patient monitoring, highlighting a wide range of patient needs^6 13 17^.

### Box 1.

Key lessons in the implementation of remote home monitoring models during the COVID-19 pandemic

- It is important to consider remote home monitoring models as an approach to maintain patients safe in the right setting.
- The use of apps for monitoring allowed the follow-up of a higher number of patients (compared to paper-based models), but some of the studies indicated that models based on telephone calls were more inclusive (i.e. including patients without internet access or technological literacy).
- Patient/carer training was identified as a key determining factor of the success of these models.
- Coordination between primary and secondary care facilitated implementation
- Primary care led models were considered, in some cases, as more adaptable to evolving patient and system needs, and easier to replicate in contexts with limited secondary care access and capacity.
- A few models integrated mental health and social care support during and after patient monitoring, highlighting a wide range of patient needs.

Despite several of the examples used apps and other types of online platforms, discussions in relation to the use of health technology were limited. The use of apps for monitoring allowed the follow-up of a higher number of patients (compared to paper-only models) but some of the studies indicated that models based on telephone calls were more inclusive (i.e. including patients without internet access or technological literacy)^19^. Patient experience was captured in some of the examples we reviewed^8 26^ but the analysis was limited. An analysis of patient experience and engagement is important as the literature on the use of remote patient monitoring for other conditions has demonstrated that higher levels of patient engagement with remote patient monitoring technology are associated with better patient outcomes^29^.

Similarly to other reviews on remote patient monitoring in other conditions, another limitation was the lack of attention placed on the implementation of the models and the failure to identify the programme theories guiding their design, factors that acted as barriers and facilitators and the extent to which the pathways were implemented according to their original plans^30^. This could be due to the limited evidence on COVID-19 and the management of patients with this disease at the time of designing and implementing these models as well as the general limited use of programme theories in the design of healthcare interventions that has already been documented in the literature^31^.

Emerging international evidence has indicated that lower thresholds for oxygen saturation, are associated with worse patient outcomes^2 32^. In the case of our review, even though some authors argued that pulse oximetry identified the need for hospitalisation when using a cut-off of 92%^16^, we could not reach conclusions in relation to patient safety and the degree to which remote home monitoring models can conclusively identify cases of deterioration at an earlier stage in the disease trajectory. The main reasons were lack of standardised reporting across articles in relation to these outcome measures and how these were measured, as well as the limitation that none of the articles used comparators.

Issues with using pulse oximetry were also highlighted such as: patient physiological measures needed to be recorded several times a day to correctly identify cases of deterioration, some remote home monitoring examples used standardised home pulse oximeters to avoid variability between different brands, pulse oximetry readings were made less accurate by nail polish, severe anaemia, hyperbilirubinemia, hemoglobinopathies, or poor peripheral perfusion from severe vasoconstriction or poor cardiac output^16 33^. Some authors also argued that patient training was a key determining factor of the success of health information technology as it ensured readings and other observations were carried out accurately^6^. Remote home monitoring needed to be seen as an approach to maintain patients safely in the right setting rather than as an admission avoidance model.

Remote home monitoring for COVID-19 patients was expected to have a positive economic impact, mainly due to costs savings in staff time and PPE utilisation, avoidance of infection of frontline medical staff and reduced hospitalisations^14 21^. However, the economic evidence in relation to these was limited. Very few of the selected studies included a simple descriptive form of economic analysis which included the cost per patient and the cost avoidances of using remote monitoring for patients with COVID-19. The selected studies have, however, raised the issue of resource allocation and funding, especially when it comes to the continuity of such programs after the first emergency situation. Most of the staff who worked on remote monitoring interventions for COVID-19 came from other services and the resources used were already existing. Yet, with the return to normal workloads, providing sufficient staff and enough resources may become a problem. Previous studies have indicated that remote monitoring in itself has contributed to increased efficiency in the use of resources (such as reduction in length of stay, increasing bed availability without compromising patient care safety, etc.)^15 21^. A complete economic analysis in this context could indicate if remote home monitoring for COVID-19 patients is a cost-effective intervention and could help inform accurate planning of the needed resources and staff. This economic analysis would also need to include costs and benefits beyond the actual remote home monitoring models, a reliable control group, as well as a longer follow up period.

This review has a series of limitations. The last search was carried on 5 February 2021, so any articles published after this date were not included. We have included preprints as a way to address delays produced by external review and publication. Furthermore, although we employed multiple broad search terms, it is possible that we missed articles that did not use these terms. Due to the variability in study designs and the descriptive nature of the articles we did not assess these for quality using standardised tools for assessment. However, we feel it is important to note that we found several cases of missing data and inconsistencies in the reporting of evaluations that would lead to low quality ratings.

The review pointed to several future areas of research. These could include an analysis of patient experience, beyond measures of satisfaction and the exploration of potential inequalities in patients’ access to remote home monitoring models or patients’ difficulties interacting with technology. Technological barriers have been reported in other studies of remote home monitoring and should not be overlooked when exploring the experiences of patients with COVID-19^34 35^. Additional attention needs to be paid to the processes used to implement these models and how these might vary based on the healthcare sector, patient population, size, wave of the pandemic and approaches used for triage, monitoring and escalation. As mentioned earlier, primary care might need to play a more central role in the coordination of remote patient monitoring models, providing more holistic care for patients and reducing the demand on hospital services^36^. The evaluation of remote home monitoring, considering its impact on patient outcomes through the use of comparators is also required. We also need to consider the sustainability of these models during multiple epidemiological peaks, compare different approaches to remote home monitoring and assess their cost-effectiveness.

## Data Availability

All of the relevant data are included in the manuscript and supplementary files.

## Contributor and guarantor information

NJF, CSJ, TG, KS, MS, SMT and CVP contributed to the design of the review. MB, KS and CVP participated in the study screening, selection and data extraction. CSJ, TG and SMT acted as cross-checkers of the extracted data. NJF, MIK, AS and KK reviewed and provided feedback on the manuscript. All authors approved the final version of the manuscript. The corresponding author attests that all listed authors meet authorship criteria and that no others meeting the criteria have been omitted.

## Dissemination declaration

The findings from this review will be disseminated widely, including to patient organisations.

## Data sharing statement

All of the relevant data are included in the manuscript and supplementary files.

## Patient and public engagement statement

Patients or the public were not involved in the design, or conduct, or reporting, or dissemination plans of our research.

## Appendix 1.

Phased search strategies

COVID-19

AND

“virtual ward” OR “remote monitoring” OR “virtual monitoring” OR “home monitoring” OR “community monitoring” OR “early monitoring”

COVID-19 OR

AND

“virtual ward” OR “remote monitoring” OR “virtual monitoring” OR “home monitoring” OR “community monitoring” OR “early monitoring” OR “pre-hospital monitoring”

AND

“silent hypoxemia” OR “pulse oximetry”

“COVID-19”[All Fields] OR “severe acute respiratory syndrome coronavirus 2”[All Fields] OR “severe acute respiratory syndrome coronavirus 2”[All Fields] OR “2019-nCoV”[All Fields] OR “SARS-CoV-2”[All Fields] OR ((“Wuhan”[All Fields] AND (“coronavirus”[MeSH Terms] OR “coronavirus”[All Fields])) AND 2020[All Fields])

AND

“virtual ward” OR “remote monitoring” OR “virtual monitoring” OR “home monitoring” OR “community monitoring” OR “early monitoring” OR “remote patient monitoring” OR “pre-hospital monitoring” OR “Covidom” OR “My m health” OR “GetWell Loop” [All Fields]

AND

“silent hypoxemia” OR “pulse oximetry” [All Fields]

**Appendix 2.**
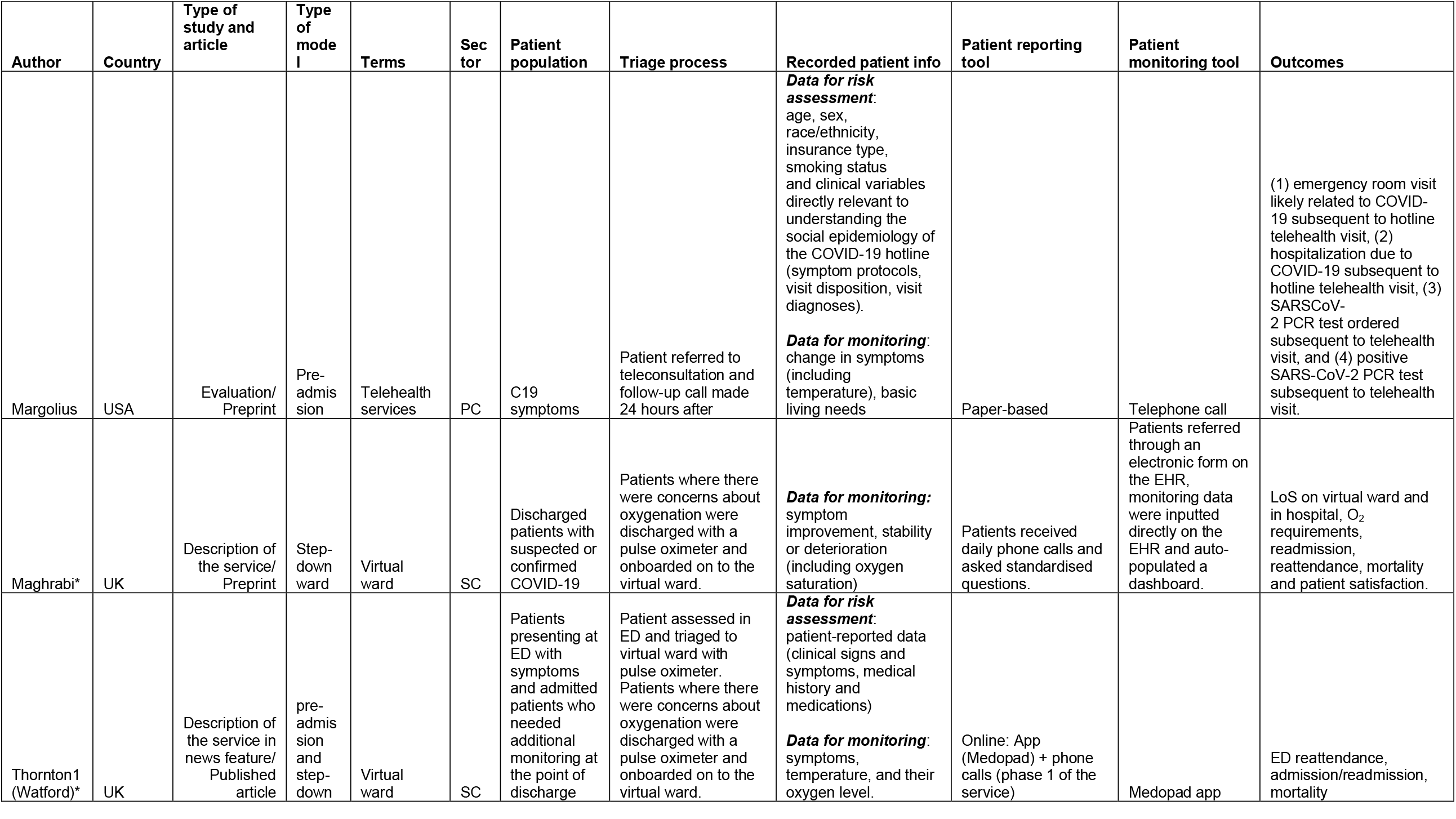
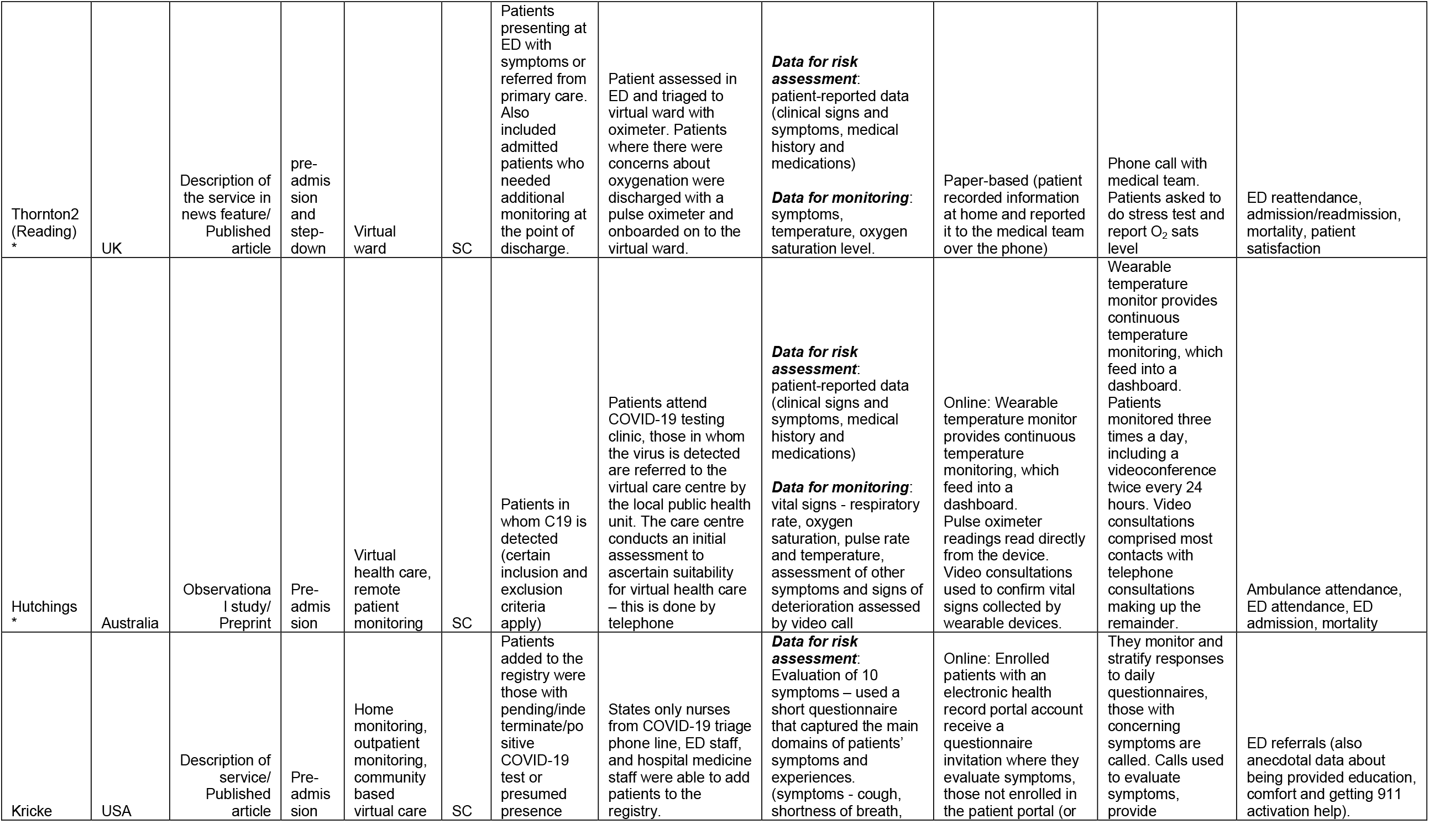
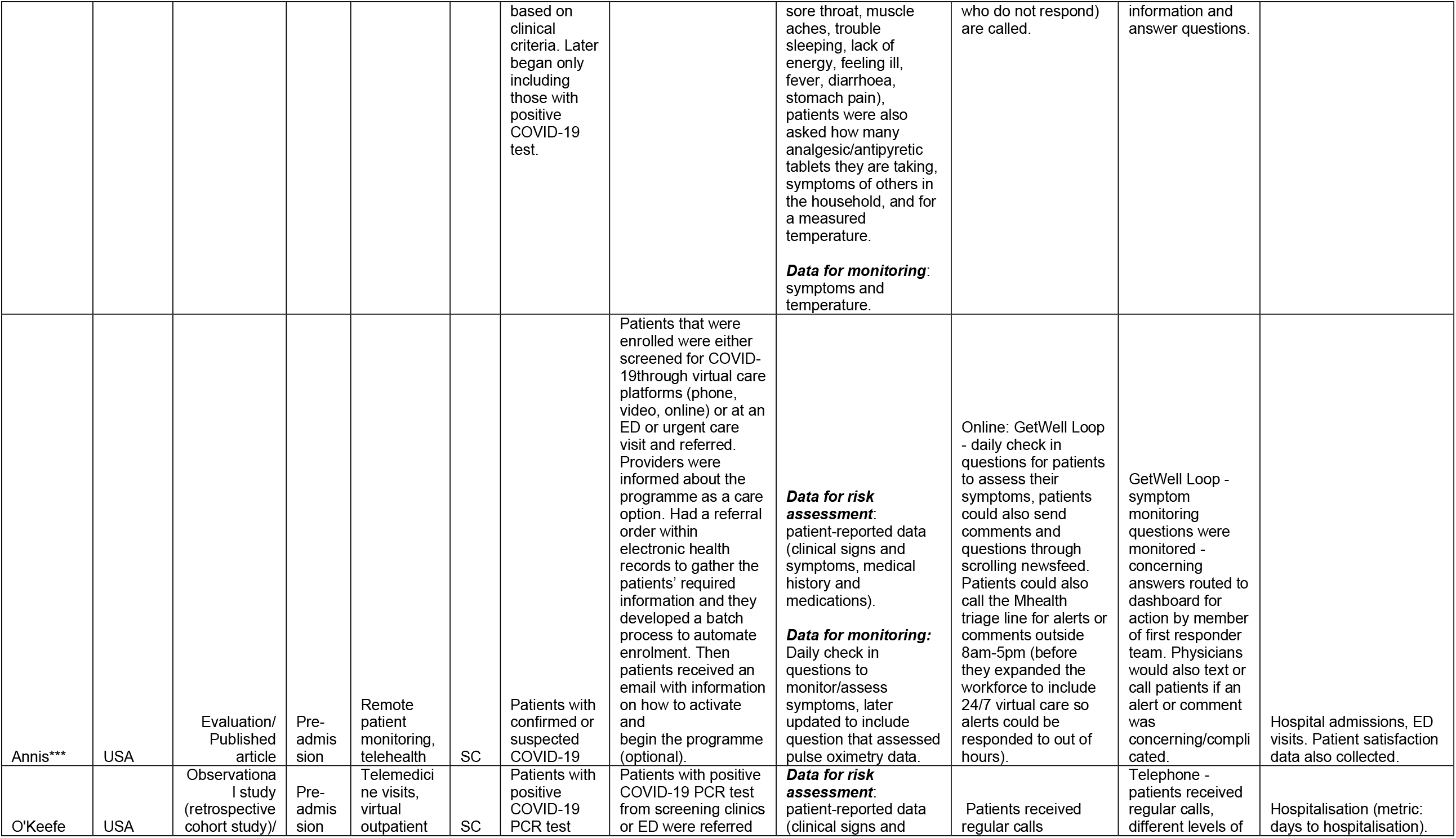
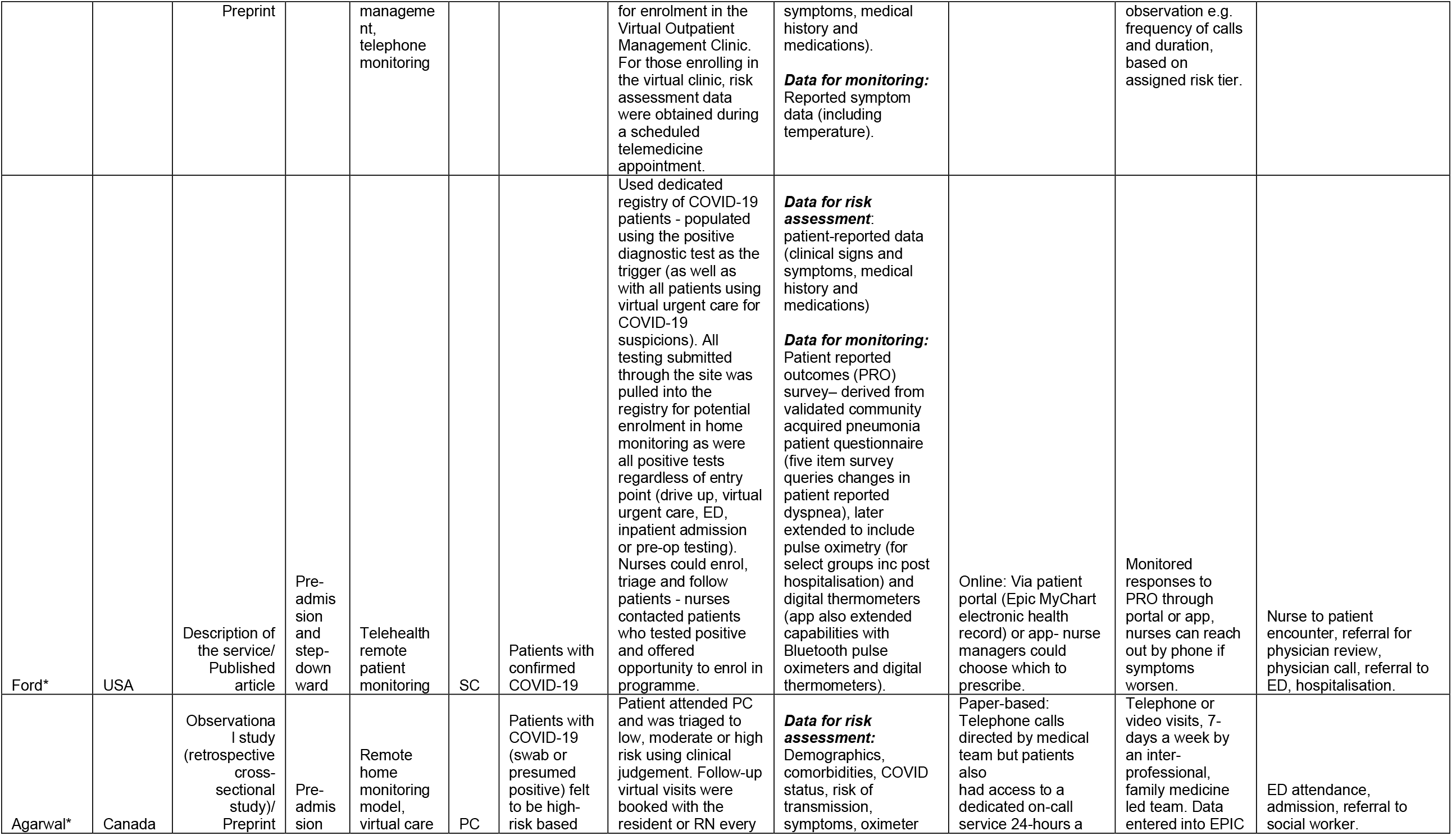
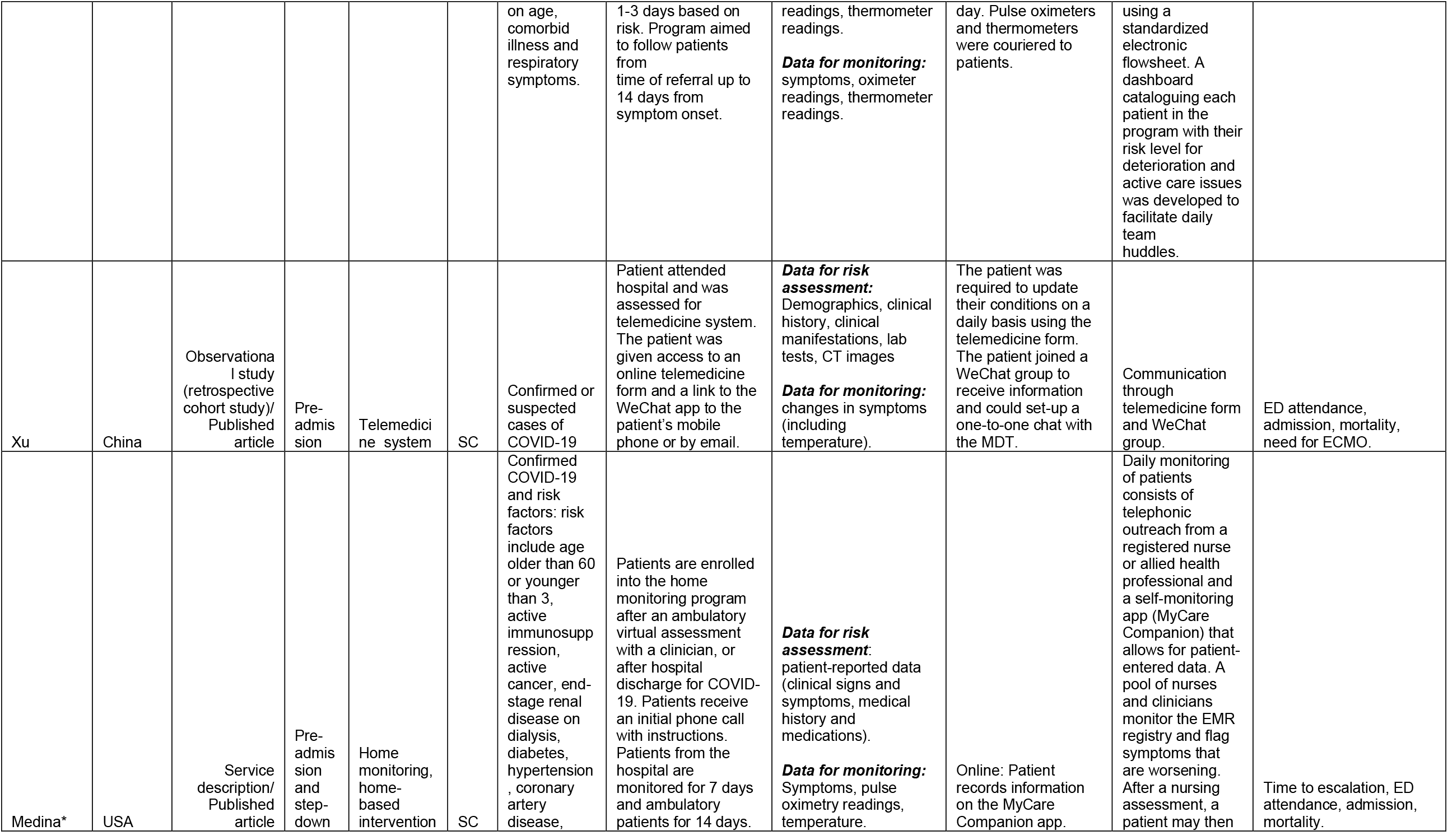
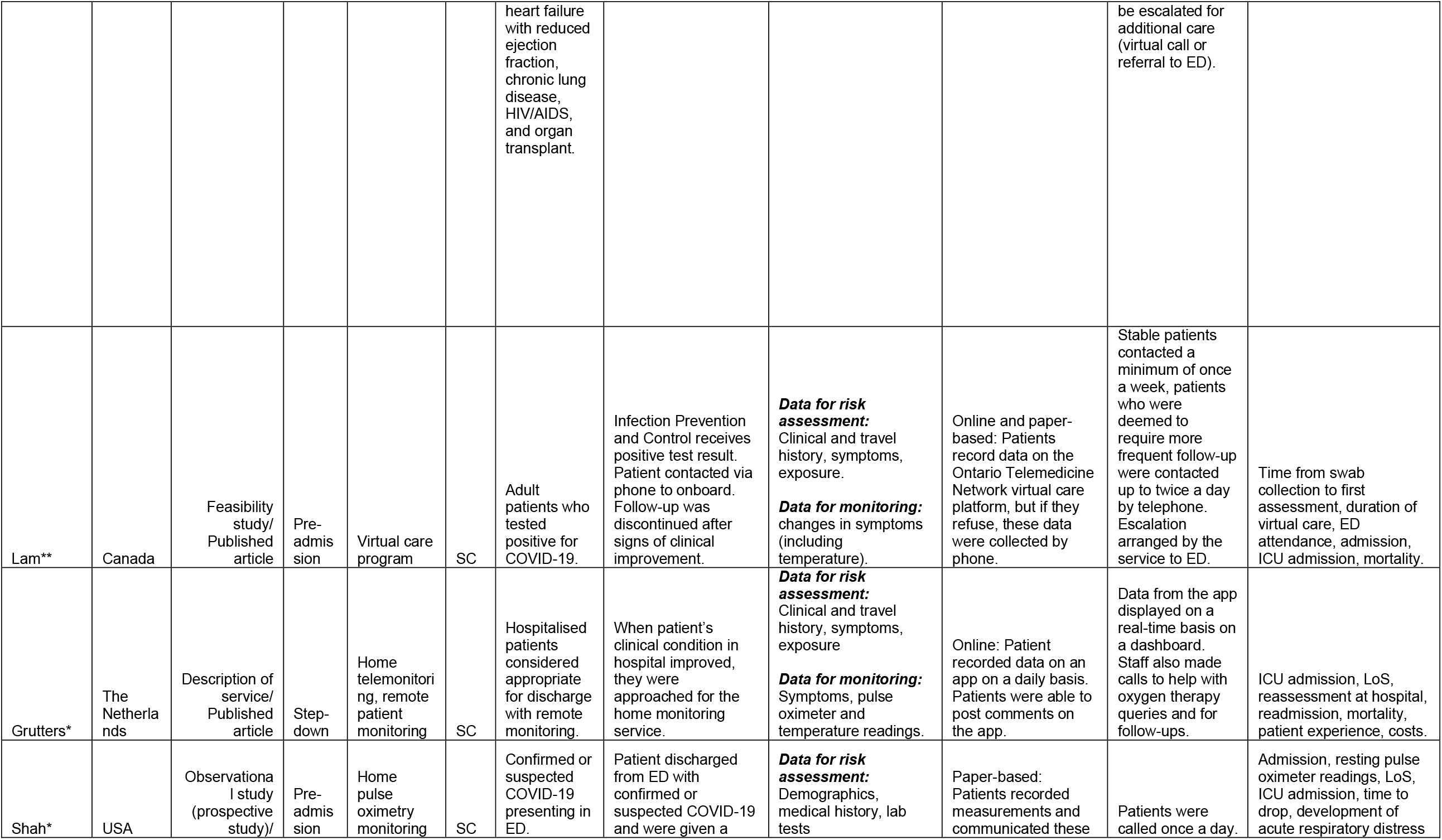
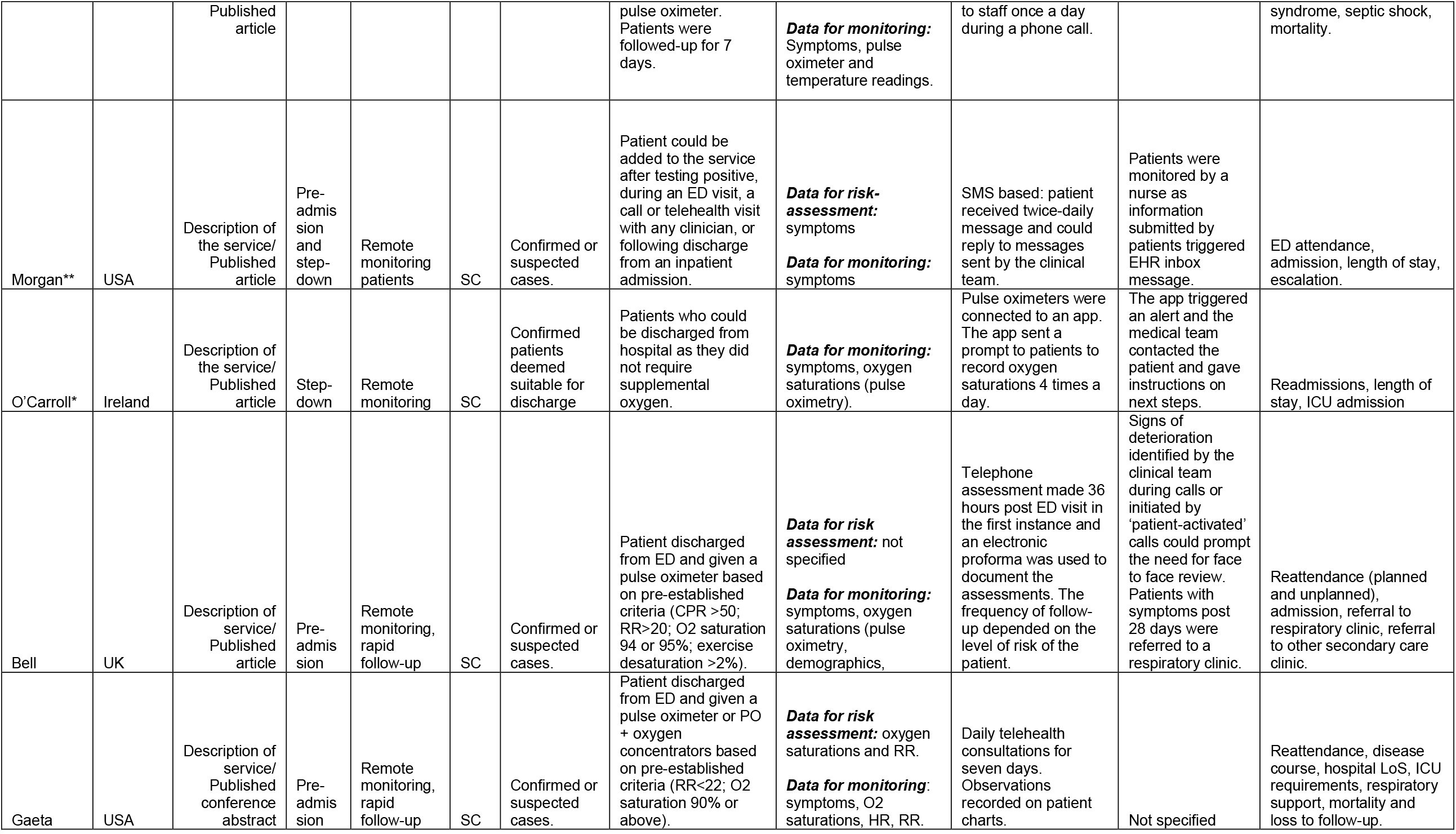
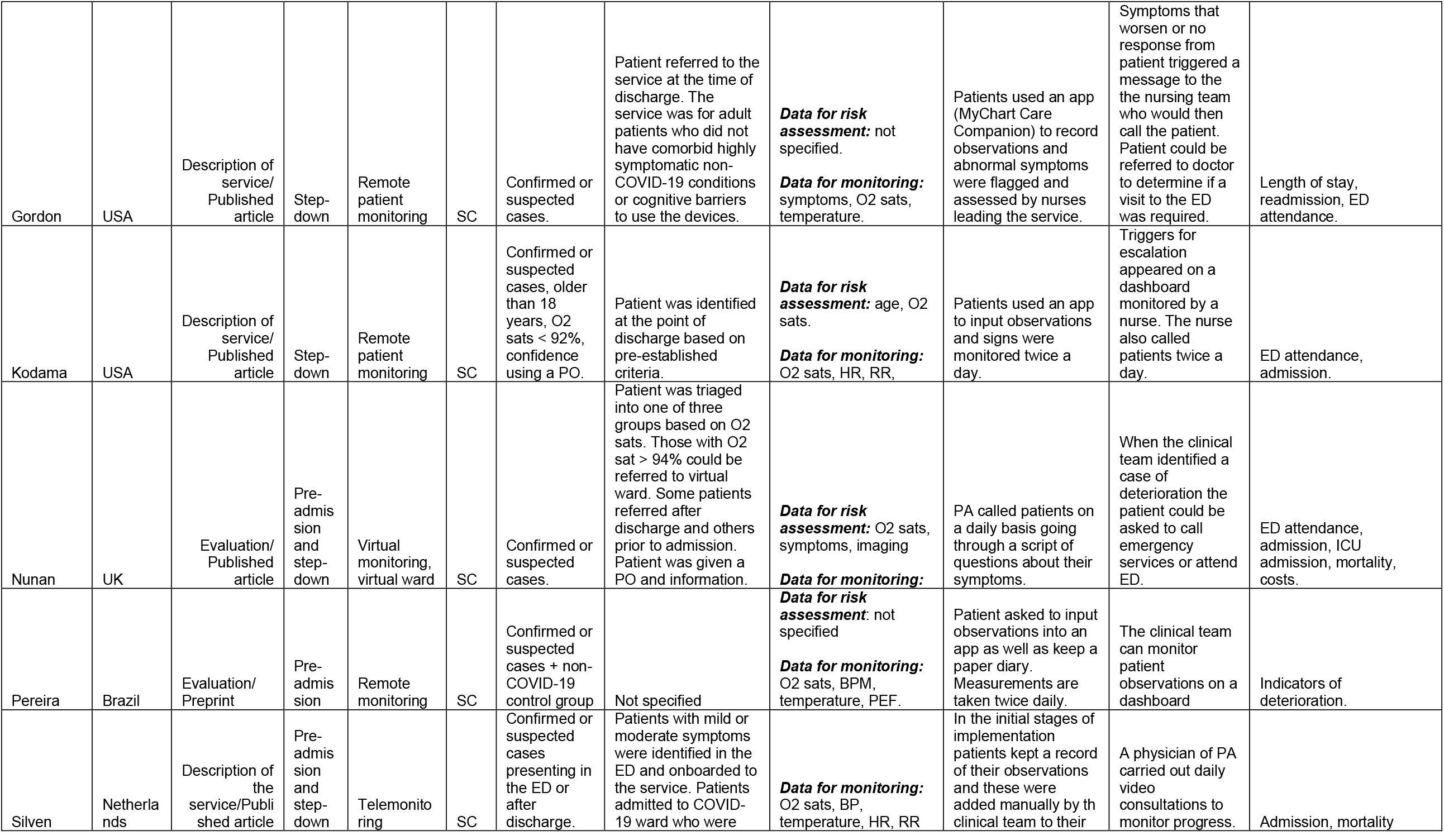
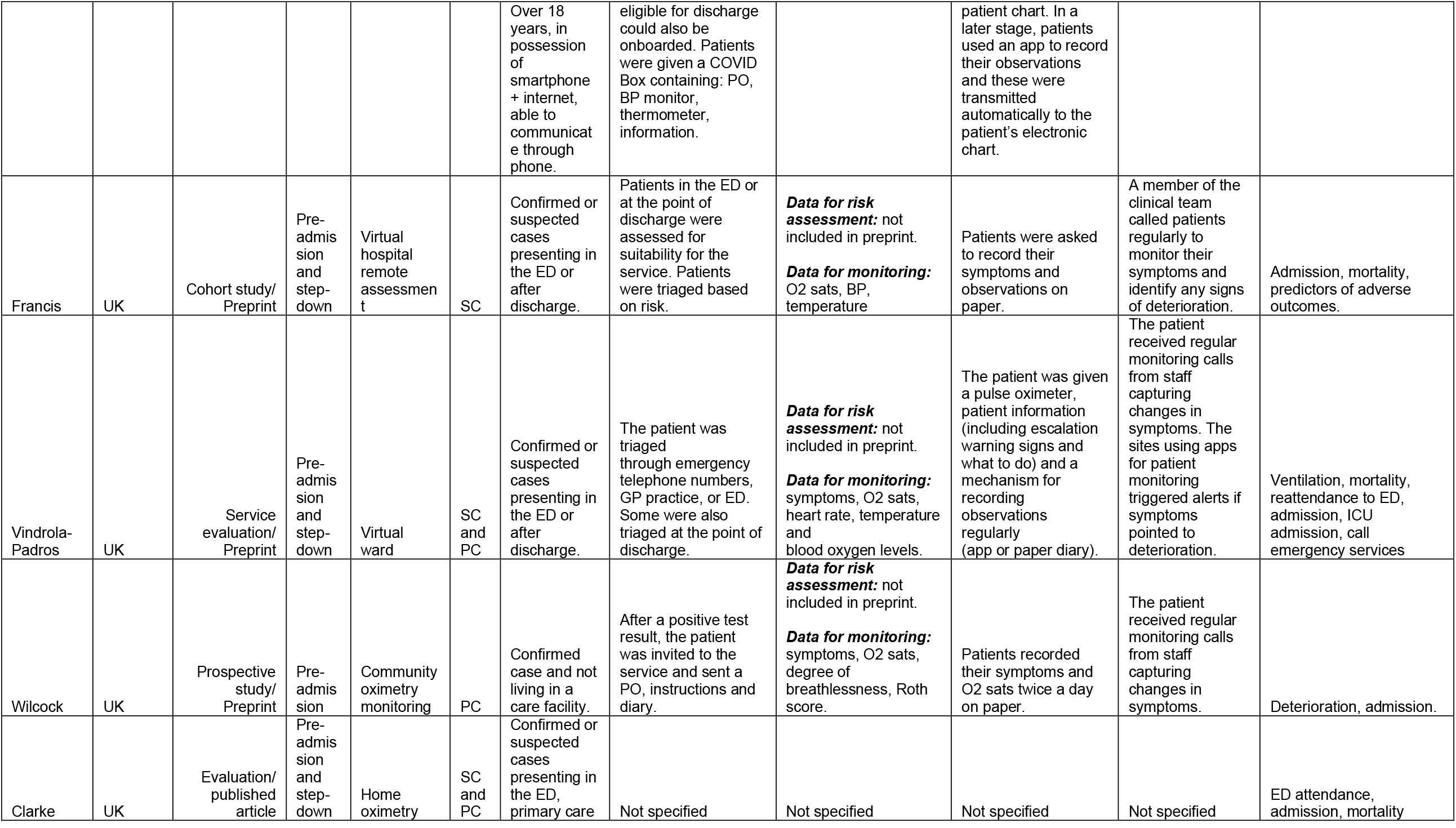
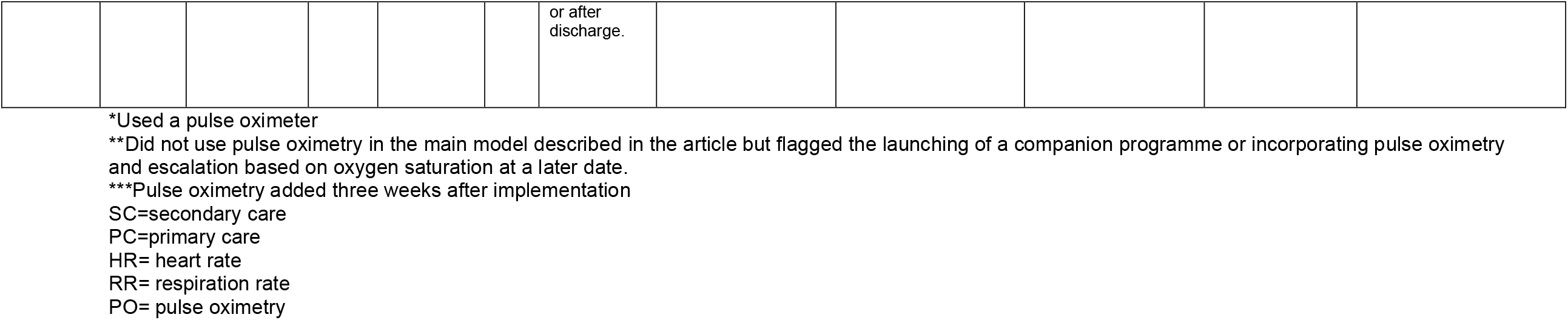
Characteristics of the included remote home monitoring examples

